# The Refined Recurrence Risk of *De Novo* variants Due to Parental Mosaicism

**DOI:** 10.1101/2025.07.02.25330538

**Authors:** Sonja Neuser, Natalie Ahmad, Denny Popp, Julia Hentschel, Johannes Lemke, Knut Krohn, Anna-Lisa Schween, Maike Karnstedt, Stephan Drukewitz, John Wiedenhöft, Rami Abou Jamra

**Author notes:** Corresponding author: Sonja Neuser, Institute of Human Genetics, University of Leipzig Medical Center, Philipp-Rosenthal-Str. 55, 04103 Leipzig, Germany. equally contributing first authors (Sonja Neuser, Natalie Ahmad). equally contributing last authors (John Wiedenhöft, Rami Abou Jamra).

## Abstract

Parents of children with genetic disorders due to *de novo* variants are counselled on a recurrence risk estimate of 1-5% for further affected siblings, while the actual probability varies between 0 and 50%. This discrepancy is well known, but barely investigated. We enrolled 135 families, in which a child had been previously identified with a pathogenic seemingly *de novo* variant (in 140 genes). Covering two germ layers, we collected blood (n=269), buccal (n=223) and nail samples (n=223) of both parents and paternal semen (n=88). Using Nanopore long read sequencing, variants were phased to the parental allele of origin. We performed deep sequencing with unique molecular identifiers of all samples. We investigated for low mosaicism with plotting identified allele fractions and Bayesian analyses. We confirmed the results individually using amplicon-based, spiked-in deep sequencing. Phasing revealed 22% of variants have occurred on the maternal and 78% on the paternal allele. Median raw target read depth achieved >7,000x, reduced to 523x after collapsing across all tissues (n≈130,000 measured values). We identified mosaicism in either the mother (n=2) or the father (n=4): four of them as mixed mosaicism, two cases as paternal gonadal mosaicism. The alternative allele fraction varied from 1.1% to 23%, while the results of the Bayesian model correlated well with amplicon-based sequencing. With 4.4%, we observe a slightly lower number of parental mosaicism compared to the literature. We now apply the amplicon-based sequencing of tissue samples – including semen – to routine counselling of parents with an affected child for individual risk assessment.

## Introduction

A significant part of severe disorders in childhood is due to *de novo* variants^1,2^. In addition to multiple challenges that the families have to deal with, uncertainties of recurrence risk complicate family planning. Empirical data suggests a general recurrence risk of 1 to 5% for having another child with the same *de novo* variant^1,3^. However, the true recurrence probability is probably much lower for the majority of families, but can reach up to 50% for a small number of them, e.g. when parental mosaicism occurs in reproductive tissues^4^. Genetic counseling to families who plan to have more children is a particular challenge for geneticists and clinicians^5,6^.

*De novo* genetic variants can arise in the parental germline or during embryonic development. The timing of these *de novo* variants’ occurrence can influence both the mutation burden and its distribution throughout the body. Some of the *de novo* variants are indeed mosaic in a parent, but affect only a small proportion of the cells. Thus, this parent shows no symptoms. Having exact information on the prevalence and distribution of the variants is a decisive factor in interpreting apparent *de novo* variants and has important implications for recurrence risk^7^. While it has become standard identifying *de novo* variants in affected children using next generation sequencing as a diagnostic tool, no sufficient technique has yet been established to detect low-level mosaicism in their parents in standard routine diagnostics. Therefore, no distinction between postzygotic *de novo* variants in the child (constitutive *de novo* variant) and postzygotic parental variants (mosaicism in a parent) can be made in daily clinical practice. Thus, medical advisors inform parents about a general recurrence risk of 1 to 5%.

Some studies used previously generated next generation sequencing data with coverage of 100x to 200x to make estimates on mosaicism in parents or in offspring^2,5,8–12^. Results gave some biologic and technical insights, but cannot be used for genetic counseling on recurrence risk.

A limited number of studies focused on specific genes. Three studies identified parental mosaicism in blood and additional tissue samples including semen in 8.6%^13^, in blood 25%, as well as in semen 17.7%^14^ and in blood 5.0%^15^ in cohorts with Dravet syndrome due to variants in *SCN1A*. Yang et al.^16^ identified parental mosaicism in blood and semen samples in 7.5% of a cohort with *ATP1A3*-associated alternating hemiplegia of childhood. Other studies were more inclusive, but had limitations. Krupp et al. analyzed a cohort of autism spectrum disorder and identified parental mosaicism in blood in 6.8%, however their analysis was based on exome sequencing data for primary mosaic identification^17^. Two studies on cases with epileptic encephalopathies identified parental mosaicism in blood and additional tissue samples without semen in 8.3%^18^ and 6.6%^19^, but with limited depth of coverage and/or number of families. Comparable numbers arise from a study on *de novo* variants in three healthy families (3.8% of all *de novo* variants are parental mosaic variants)^4^.

More diverse cohorts with neurodevelopment delay disorders identified parental mosaicism in blood in 3.0% (14/237 trios)^20^, in blood or semen in approximately 6.8% (3/44 trios)^21^ and in blood and several tissue samples including semen in 10.2% (6/59 trios)^10^.

As the available studies describe small cohorts, only test blood samples, or are highly specialized to few genes, gaining an overview of the available data and developing solid methods for identifying parental mosaicism is crucial for a solid genetic counseling.

In this study, we aim at generating reliable insights into the development and representation of *de novo* variants in different parental tissues of a broad spectrum of severe early onset disorders. We develop a laboratory procedure and a clinical recommendation to determine the personalized recurrence risk for future siblings of children with a *de novo* variant.

## Subjects and Methods

### Selection of families, recruiting and sample collection process

Out of a cohort of approximately 7,000 next generation sequencing analyses at the Institute of Human Genetics between 2016 and 2021, with a broad spectrum of disorders, we identified index cases with an apparent *de novo* variant. We focused on young individuals (born 1996 or later) in order to increase the chance of their parents’ interest and availability for research. We contacted approximately 300 families. The Ethical Committee of the Medical Faculty, Leipzig University, approved genetic testing in a research setting for all probands (014/19-ek). Written informed consent of the parents to publish genetic and clinical data was received and archived by the authors.

Eventually, 135 families were included. We analyzed 137 clinically relevant variants (in three families two clinically relevant *de novo* variants were identified, two children carried the same *de novo* variant). Following their informed consent, samples of the families were collected. In addition, we reviewed 25 available trio exome datasets of the above-mentioned cohort for *de novo* variants in non-disease-associated genes and added 39 *de novo* variants to the analyses.

### DNA extraction, deep sequencing with unique molecular identifiers (UMI)

DNA extraction of buccal and EDTA blood samples was performed using the MagCore 101 Kit. Blood filter cards were reduced in size with sterile scissors and dissolved in GT Buffer solution before the enzyme solution was added. Afterwards, MagCore 401 Kit was used to extract the DNA. Semen samples were incubated for one hour after the initial addition of GT buffer, 1M DTT and Proteinase K enzyme solution to the samples. After that, DNA was isolated using the MagCore 401 Kit. DNA extraction from finger nails was done using the DNeasy® Blood & Tissue Kit from Qiagen. First, nails were either crushed mechanically or cut into smaller fragments with sterile scissors before flushing multiple times with H_2_O. Then, they were mixed with Lysis Solution TLS buffer, Proteinase K and 1M DTT buffer solutions to prepare them for two hours of incubation. Afterwards, samples were pipetted onto specific membranes with DNA absorbing abilities together with buffer solution and washed multiple times. After several centrifugation and incubation periods, the DNA was eluted in the provided buffer.

DNA quality and concentration control was performed using NanoDrop 2000™ or Qubit™ 3.0.

We performed targeted deep sequencing on all samples, using a custom panel from TWIST Bioscience (TWIST Bioscience, San Francisco, CA, USA; target regions of panel in Table S1 in Zenodo^22^, sheet “target regions panel”). In addition, unique molecular identifiers (UMI) were used to identify PCR duplicates (IDT xGen Duplex Seq Adapters; IDT Integrated DNA technologies, Coralville, IA, USA). The goal of using UMI was to reduce false positives in very low fraction variants and allow for the detection of the correct variant allele fraction (VAF) through identification of amplification errors. Wetlab processing including enrichment and library preparation was performed according to manufacturer’s instructions. Sequencing was done on NextSeq500 (Illumina, SanDiego, CA, USA).

### Bioinformatics

We implemented the bioinformatics pipeline to create input for our statistical model in SnakeMake (7.32.4)^23^. All computations were done in a virtual Conda (24.5.0)^24^ environment using Mamba (1.5.8)^25^. We used fastp (0.23.4)^26^ for QC and UMI trimming of reads in FASTQ files. Reads were mapped using BWA (0.7.17)^27^ and merged using samtools (1.19.2)^28^. UMI groups were created using fgbio (2.3.0)^29^. Filtering of reads overlapping target regions was done using pysam (0.22.0)^30^. Consensus was computed using fgbio^29^.

For auxiliary steps in the pipeline such as indexing, sorting etc., we used samtools (1.19.2)^28^, bedtools (2.31.1)^31^, gawk (5.3.0)^32^, R with tidyverse (2.0.0)^33^, htslib (1.19.1)^28^, NumPy (1.26.4)^34^ and Picard (3.1.1)^35^. Naïve calling of variants was done using vardict with a minimal VAF of 0.001 and a minimum read count of 2^36^.

### Statistical model

We designed a Bayesian model for allele fractions and global error rate as follows: Let *v* denote a genetic variant, *r* a parent, and *t* a tissue type. Let *ref*(*v*) and *alt*(*v*) denote the DNA sequence of reference and alternative alleles of variant *v*, respectively; we assumed no third allele to be present in either parent or child. Let *par*(*v,r,t,d*) denote the true allele sequence in DNA template *d*, and *seq*(*v,r,t,d*) its observed sequence in the NGS data set; due to the use of UMIs, we assumed that no duplicate sequencing reads existed for any template *d*. Let *L_ref_*(*v,r,t,d*) and *L_alt_(v,r,t,d*) denote the Levenshtein distance of *seq*(*v,r,t,d*) to *ref*(*v*) and *alt*(*v*), respectively. As all child variants have been confirmed using Sanger sequencing, we assumed *alt*(*v*) to be reliable and do not model potential discrepancies between the true and observed allele sequences in the child. Let *vaf*(*v,r,t*) denote the VAF of variant *v* in tissue *t* of parent *r* if *v* is a pathogenic variant affecting their child, and constant 0 otherwise (as the variants are pathogenic and practically absent in the general population (gnomAD), we assumed *a priori* that parents do not carry any variants of interest affecting children unrelated to them). Let ε be the global per-base error rate (deletion, insertion, substitution) across all reads due to the experimental pipeline. To obtain posterior distributions for ε and *vaf*, we derived the likelihood function terms as follows (indices have been dropped for readability):

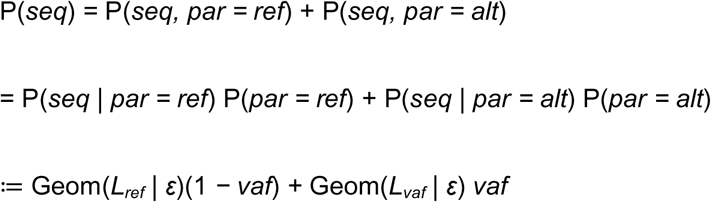

The geometric distribution over the Levenshtein distance models the minimum number of accumulated single-base experimental errors required to transform the true sequence *par* into the observed sequence *seq* (note that more complex error distributions could be substituted here without affecting the structure of the model). We assumed Jeffreys priors *vaf* ∼ Beta(½, ½) and (1 − ε) ∼ Beta(½, ½) for the latent parameters, to reflect absence of prior knowledge due to the foundational nature of this work, and to ensure reparametrization invariance. Posterior distributions were summarized using maximum a-posteriori (MAP) estimates and high-density intervals (HDI, convex closure of high-density regions).

In total, our data comprised 811 parent/tissue combinations, yielding 1,058 latent variables for VAF (due to multiple variants in some families) and over 279 million observations of tuples (*L_ref_*, *L_alt_*). We implemented our model in Stan using the CmdStanR interface (version 0.8.0). As the definition of *vaf =* 0 for unrelated individuals yields an analytical solution for the beta posterior due to its conjugacy with respect to the geometric distribution, posterior hyperparameters for ε were computed directly for those cases and passed to the prior in the model. Posterior inference of *vaf* and ε on the remaining ∼2 million tuples was then completed using 40 parallel MCMC chains of the No-U-Turn Sampler (NUTS), each running for 2000 iterations. This two-step procedure, mathematically equivalent to full sampling, yielded a running time of only 2.4 minutes, a 34.9-fold speedup compared to 1.4 hours for full sampling. Each parameter exhibited an RLJ ≈ 1 (all <1.003), indicating excellent mixing diagnostics. Bulk and Tail Effective Sample Sizes were between 13,926 and 69,782, and hence well above the recommended minimum of 100, indicating highly reliable posterior quantiles.

### Replication with different methods

In order to replicate our results, we validated seven mosaicism samples (paternal semen sample of IND_027; maternal blood and buccal sample of IND_038; paternal blood, buccal and semen sample of IND_051; paternal semen sample of IND_097) and nine control sample (Table S1 in Zenodo^22^, sheet “Primer_validation”). We amplified the specific variant region with regular primers (available from segregation analysis with Sanger sequencing; Table S1 in Zenodo^22^, sheet “Primer_validation”). We then spiked-in the PCR products in an NGS run.

### Long-range PCR, long-read sequencing and phasing

In order to identify the origin of the specific *de novo* variant, we used variant-specific long-range PCR and subsequent long-read sequencing of the products on a Nanopore device. Primers were designed using Primer3 as a batch with a product size range of 9-10 kbp around the variant of interest. Two primer pairs were chosen manually, and in case of insufficient results, we designed additional primers (Table S1 in Zenodo^22^, sheet “primer_phasing”).

We amplified DNA from both parental blood samples and the proband’s sample using GoTaq® Long PCR Master Mix by Promega. After preparing the samples, we performed the PCR for 30 cycles with ligation temperature of 65°C, according to manufacturers’ instructions.

PCR products were pooled and subjected to library preparation using the NEBNext Companion Module (New England BioLabs) and Ligation Sequencing Kit (SQK-LSK109, SQK-LSK110 or SQK-LSK114, Oxford Nanopore Technologies) according to the standard protocol with the following adjustments: Incubation time of the end-prep reaction was increased to 30 min at 20°C and 30 min at 65°C. Incubation time of adapter ligation reaction was adjusted to 30 min at room temperature. Individual samples were not barcoded, but sequenced separately if the same primer pair was used for more than one family. Furthermore, family members were sequenced separately. The libraries were loaded onto Flongle or MinION flow cells of type R9.4.1 or R10.4.1 and sequenced on a MinION device Mk1b (Oxford Nanopore Technologies). Raw sequencing data (fast5 or pod5 files) was base-called and mapped against the reference human genome GRCh37 using guppy (Oxford Nanopore Technologies) and a super accuracy base-calling model with enabled read splitting, adapter trimming and calibration strand removal.

Reads in the target region were manually inspected in IGV (latest version up to version 2.16.2) for informative SNPs. Different options included: heterozygous or homozygous SNP in one parent and in *cis* (within the same read) with the *de novo* variant, homozygous SNP in one parent and not within the same read of the *de novo* variant or more complex phasing analyses with multiple SNP haplotypes. In trios with inconclusive haplotype analyses, we also used read quality and read sorting to visualize more complex SNP haplotypes.

### Literature overview

To establish an overview of the state of literature and perform a meta-analysis, we have identified relevant studies through a comprehensive PubMed search using the keywords “parental mosaicism *de novo* variant” (last search: 22^nd^ of June 2025) and cross searching through references of genetic research papers on the subject. Case reports were excluded. We collected 29 relevant publications between 2014 and 2025, and systematically extracted core data including the cohort size, cohort composition, applied methods, NGS coverage, mosaicism in blood and total mosaicism in different tissues (in percentage). Twenty studies were excluded from further analysis because of different variant type, different approach of identifying parental mosaicism, postzygotic mosaicism in affected individual, specific genes with known higher or lower number of parental mosaicism or known remarkable family history. Details are in Table S2 in Zenodo^22^.

To obtain pooled estimates of mosaicism, we calculated the weighted mean and variance using cohort size as weights. The weighted mean and variance were computed using the Hmisc package in R. The effective sample (n_eff_) size was calculated based on cohort size per study. The standard error was derived from the weighted standard deviation divided by √n_eff_, and the 95% confidence interval was computed using the t-distribution with (n_eff_-1) degrees of freedom.

## Results

### Cohort, samples, and variant spectrum

The age of the affected persons at the time of setting a diagnosis varied between two months and 31 years with a median of 4 years (mean 5.9 ± 5.6). The time between diagnosis and enrollment in the study was on average 21.0 ± 14.9 (median: 19) months. Most children have developmental delay (113 of the children have an HPO term of Neurodevelopmental delay: HP:0012758 or its downstream branches). Median age of mothers at conception was 31 years (mean 31.0 ± 5.1) and fathers 34 years (34.2 ± 6.4). Upon enrollment, the median age of mothers was 40 years (mean 39.8 ± 6.8) and fathers 43 years (mean 43.1 ± 8.1). This also represents the age of the fathers at time of semen sample collection (Figure 1C and 1D and Table S1 in Zenodo^22^, sheet “clinical” for details of the cohort).

**Figure 1.**
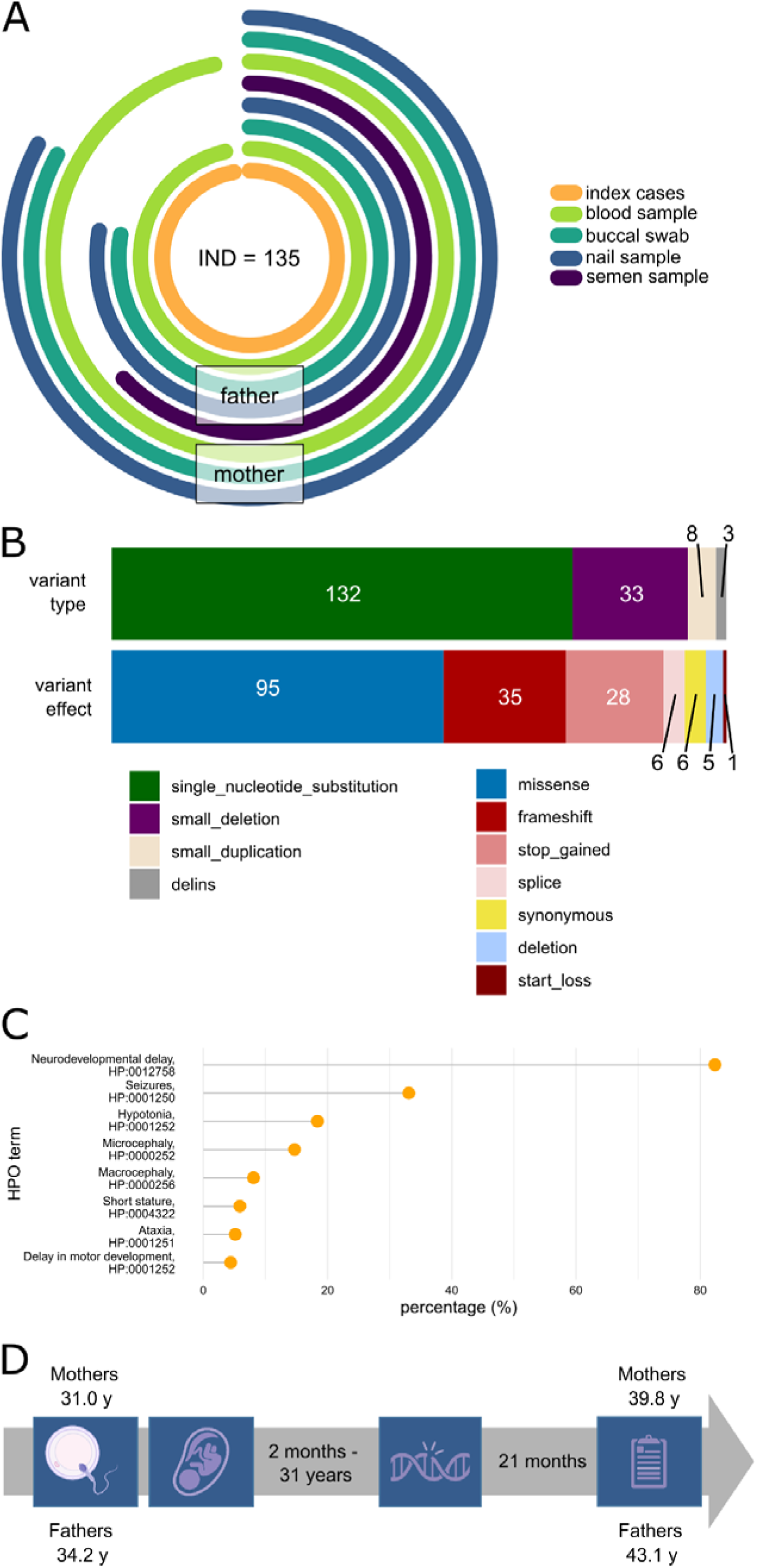
A. The cohort of 135 families with 135 affected children compiles 269 blood samples (green), 223 buccal swabs (light blue), 223 nail samples (dark blue) of the parents and 88 semen samples (purple) of the fathers. B. Distribution of variant type and variant effect of all analyzed 176 de novo variants. C. Most frequent HPO terms of the affected children (most children are documented with more than one HPO term). D. Schematic depiction of chronology of the study with mean age of the parents at conception of the child, age of the child at genetic diagnosis, mean time between diagnosis and study enrollment and mean age of the parents at study

Covering the ectodermal and mesodermal embryonic germ layers, we collected EDTA blood or blood filter cards (n=269), buccal swabs (n=223) and finger nail samples (n=223) of both parents and a semen sample of the father for direct germline testing (n=88) (Figure 1a). During the course of the study, one sibling (of IND_027) was diagnosed with the same pathogenic variant in *NFIX* of the index via targeted Sanger sequencing following clinically suspected Malan syndrome. The sibling was not enrolled as part of the cohort.

Taken together, 137 pathogenic variants in 102 different genes were investigated; one variant in *SHANK3* (VAR_02; c.3679dupG, p.(Ala1227Glyfs*69)) was identified in two unrelated children (IND_023 and IND_029). In three children (IND_034, IND_057 and IND_103), two different pathogenic variants as dual diagnoses were found (Table S1 in Zenodo^22^, sheet “variants”). The genes, in which the highest number of distinct variants occurred in the cohort, were: *ARID1B* (n=5), *MED13L* (n=5), *SCN1A* (n=4), *KMT2A* (n=4). We included further 39 *de novo*, clinically irrelevant variants that we identified in 41 available trio exome datasets of the cohort. Overall, we included 176 variants, of these 132 single nucleotide substitutions, 33 small deletions, 8 small duplications and 3 delins (Figure 1b).

### UMI-sequencing

Uncollapsed reads of deep sequencing reached a median raw target read depth of >7,000x across all samples. After grouping and consensus calling, the final median target read depth after UMI processing was 523x (25th percentile: 329x, 75th percentile 811x, allowing a median detection rate of 0.1% in the naïve VAF). Plots of VAF >0.2% of all variants and all samples after UMI collapsing is in Figure 2.

**Figure 2.**
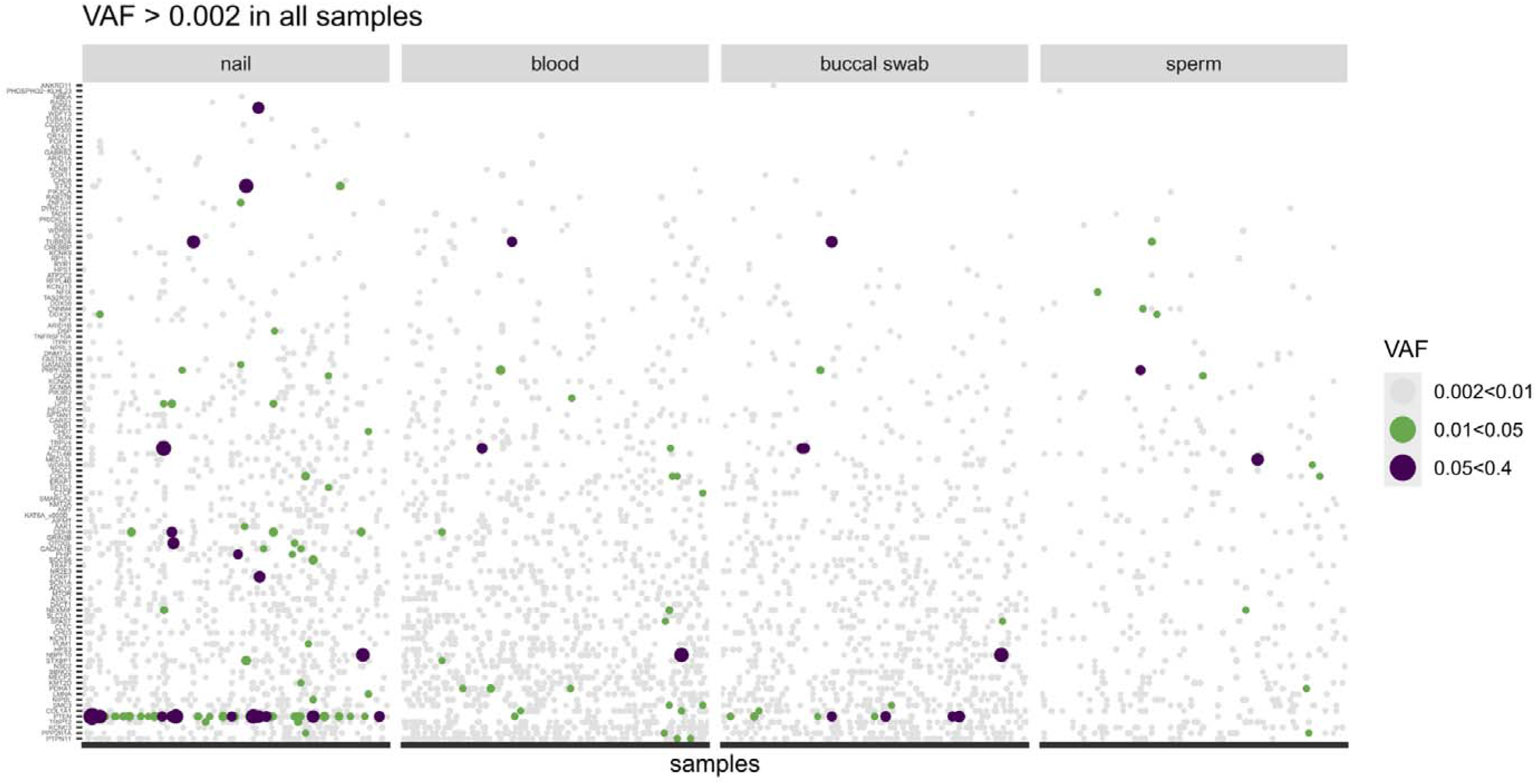
Naïve VAF after UMI collapsing: plot shows that background noise is a common phenomenon. Thus, we moved to Bayesian model to calculate most probable VAF. See below.

### Mosaicism

Naïve counting of alleles and reads and consideration of allele fractions led in both uncollapsed and collapsed datasets to a large number of obvious artifacts and a low signal-to-noise ratio (Figure 2). Thus, to achieve reliable results on mosaicism and separate false positive hits from low-grade mosaicism, we developed a custom-designed Bayesian VAF model, which also explicitly computes a posterior distribution for the experimental noise (see below).

We identified mosaicism in six families (6/135 families = 4.4%; 6/176 variants = 3.4%). Four cases are of disease-associated variants, two cases are of the additional, clinically irrelevant *de novo* variants. Paternal mosaicism of the four collected tissues blood, nails, semen, and buccal swab (mixed mosaicism) for two fathers (IND_051, IND_046), paternal mosaicism only in semen was identified in two fathers (IND_027, IND_097). Mixed maternal mosaicism in the three collected samples was found in two mothers (IND_038, IND_122). No isolated mosaicism of one tissue was found in mothers. In addition, we found no fathers that have mosaicism in blood, nails, or buccal swab but not in semen.

VAF range with UMI collapsing in the mixed mosaicism parents was between 1.1% and 22.7%, in both fathers with gonadal mosaicism VAF was 1.5% (IND_027) and 12.8% (IND_097) (Table in main text and Figure 3 a and b). Maximum a posteriori (MAP) estimates of VAF are between 0.7% and 22.5% in the mixed mosaicism parents, and were 1.0% and 12.5% in both fathers with gonadal mosaicism. The maximum a-posteriori estimate for the total experimental error rate ε was very low at about 0.439%, likely due to corrections achieved by UMI consensus.

**Figure 3.**
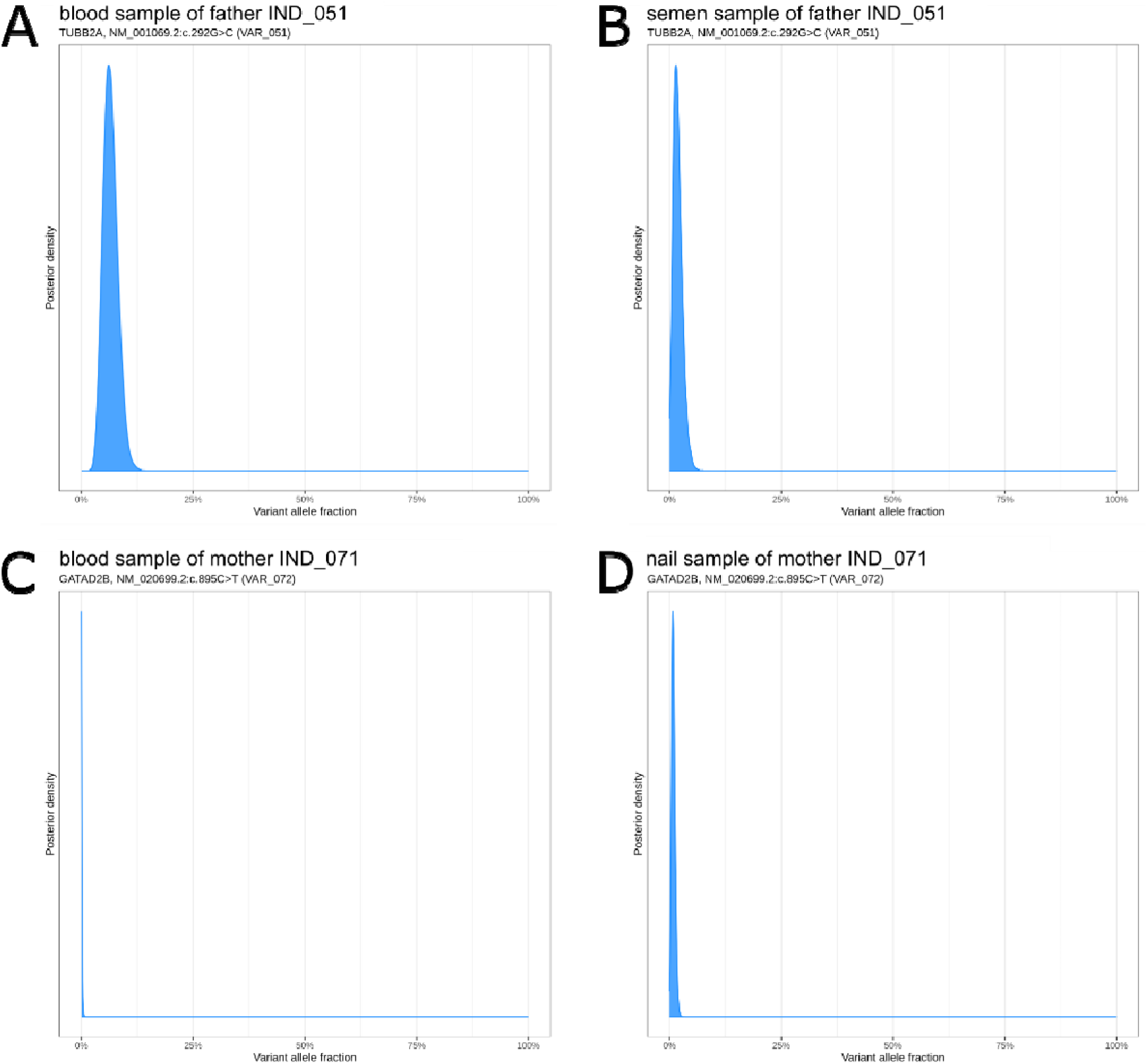
A. Example of a mosaic for the variant VAR_051 in the blood sample of the father of IND_051 with a maximum a posteriori probability (MAP) of 0.065. B. Example of the same variant VAR_051 in the semen sample of the father of IND_051 with a MAP of 0.015. C. Example of no mosaic for the variant VAR_072 in the blood sample of the mother of IND_071 with a MAP of 9.90E-06. D. Example of a suspected contamination for the variant VAR_072 in the nail sample of the mother of IND_071 with a MAP of 0.008.

### Validation with different method

With a mean coverage of 70,500x of the NGS-spike in amplicons, we received comparable results regarding the alternative allele fraction of the specific variant (Table 1). This indicates that performing an individualized approach with NGS-spike in amplicons is possible and would allow a search for low-level mosaicism in the parents in a diagnostic setting.

**Table 1.**
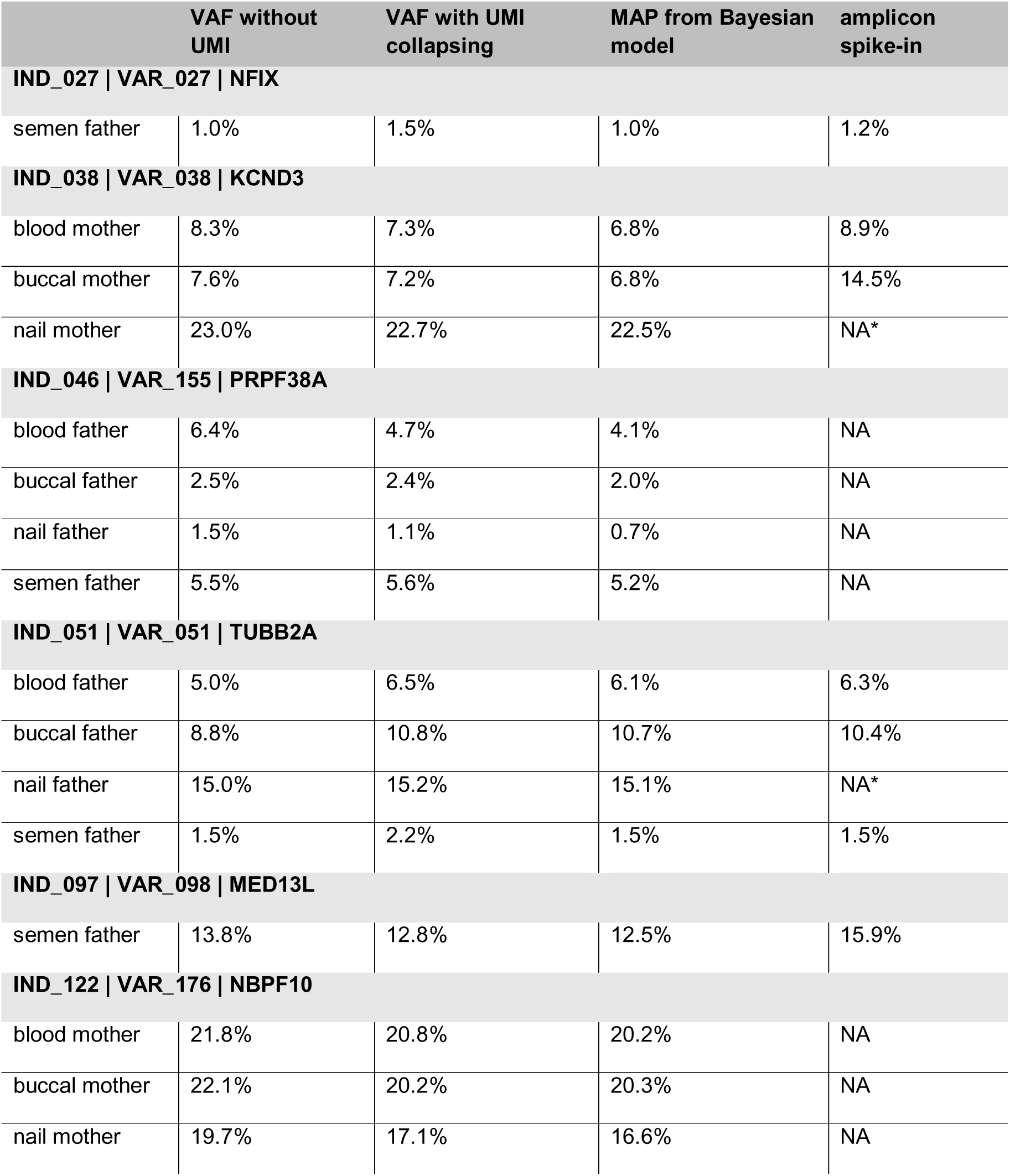
Identified parental mosaic variants with comparison of variant allele fractions (VAF) from different analysis approaches. First and second column with VAF results from two different bioinformatics pipelines (with or without UMI collapsing) using naïve counting. Third column with results from the Bayesian model given in maximum a posteriori probability (MAP) of VAF. Fourth column with results of validation of previously confirmed mosaicism using amplicon spike-in. Comparable results allow an individualized approach and a search for low-level mosaicism in the parents in a diagnostic setting using NGS-spike in amplicons. NA: data not available (not performed or *sample depleted)

### Phasing

As a first stratification, long-read sequencing and phasing was successful for 81 variants (46%). 63 were phased on the paternal allele (78%) while 18 variants were phased on the maternal allele (22%). In most unphased cases, the long-range PCR was not successful (with too low coverage of long-read sequencing) after two sequencing rounds for phasing analysis. In further cases, we could not identify an informative haplotype within the long-range fragment for phasing or variants were not accessible with long-read phasing due to the local genomic characteristics or variant type (homopolymeric region or larger deletions) (Figure 4 A and B) or for other lab-specific reasons.

**Figure 4.**
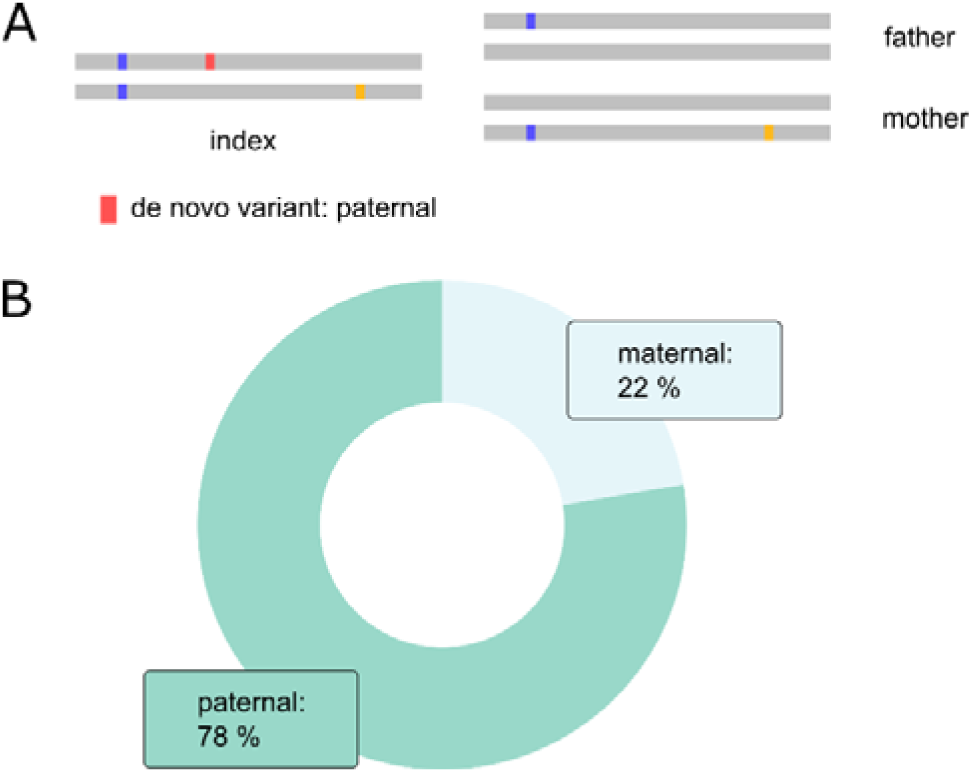
A. Schematic depiction of phasing a de novo variant using long-range PCR amplicons. With at least one other heterozygous variant, we can haplotype the de novo variant of interest. B. Percentage of maternal or paternal origin of the de novo variants after phasing with long-read sequencing.

### Literature overview

We used the data of nine publications for a weighted meta-analysis based on literature review^4,18–21,37,10,38,39^. These studies gathered comparable collected cohorts of in total 1,116 families with a size between 44 and 264 family trios with a diverse spectrum of genes. Four cohorts compiled only blood samples and five cohorts compiled additional tissue samples (oral mucosa, urothelium, semen). Seven of these used deep NGS with either targeted amplicons or MIPs for identification of mosaic variants, while two studies used ddPCR (Table S2 in Zenodo^22^).

The identified rate of mosaicism for a mosaic for the *de novo* variant in one parent ranged from 2.3% to 15.0% with a weighted mean of 4.7% (95% CI: 2.4-7.0) in blood and from 6.6% to 20.0% with a weighted mean of 8.8% (95% CI 3.6-14.0) in all tissue samples. Overall weighted mean together with our study was 7.5% (95% CI 3.0-11.9) in all tissue samples

## Discussion

The rate of germline mosaicism among parents in our cohort is 4.4% in all analyzed tissue samples. The VAF shows considerable variability across different tissues in both parents. Notably, blood and buccal swab samples demonstrate better consistency (Pearson’s r = 0.95) than blood and nail samples (r = 0.46). Buccal swabs may contain up to 25% leukocytes, which originate from the mesodermal embryonic layer^40^. Statement on the consistency of blood and semen samples is limited based on only two data points. But with 1/3 of mosaic cases with isolated gonadal mosaicism, our findings suggest that comprehensive tissue-specific testing is necessary to achieve accurate results.

We identified a slightly lower number of paternal mosaicism in comparison to the synopsis of nine international, comparable studies. Potential explanations for the lower mosaicism rates are

– Reduced sensitivity of our testing methods. However, the mosaicism reported in the six studies generally exhibits VAFs detectable by our approach. Our testing identifies mosaicism at rates of 0.2% before UMI collapsing and 0.3% afterward, aligning with the detection thresholds of other comparable methodologies.
– Other studies may have considered background noise as mosaicism as in some cases there is no independent validation of the identified mosaicism and no clear picture on the level of noise that is associated with the used methods (compared to our Figure 2).
– As we recruited our cohort, we were fully agnostic to the family history. Thus, we do not have a bias for recruiting families with known/expected mosaicism due to family history or the affected gene.
– Coincidental variabilities due to relatively small cohort sizes of all studies

Certain genes are predisposed to gain-of-function missense variants that undergo positive selection in testicular tissue, providing a clonal proliferation advantage as men age. These “selfish sperm” variants are well-documented in genes involved in the RTK-RAS-MAPK signaling pathways, including *FGFR2, FGFR3, HRAS, PTPN11, RET*, and *SMAD4*^41,42^. In our cohort, we identified one individual carrying a *PTPN11* variant (VAR_024). Phasing confirmed - as expected - paternal origin of this *de novo* variant. Despite the potential for a semen-specific mosaic pattern often associated with these variants, we did not detect such a mosaicism. Our findings emphasize that while some genes exhibit well-characterized *de novo* mutational patterns, such as selfish selection, these phenomena do not universally explain all cases. This highlights the necessity of conducting individualized analyses to accurately assess recurrence risks and variant origins on a case-by-case basis.

According to the literature, additional genes have been identified that exhibit higher recurrence risks for seemingly *de novo* variants, suggesting potential parental mosaicism. In our cohort, one family (family of IND_027) presents with two children affected by a variant in *NFIX*, despite the father’s semen sample showing a very low VAF. Variants in *NFIX* are well-documented in the context of Malan syndrome, with reports of sibling recurrence attributed to parental mosaicism^43–45^. However, systematic analyses of recurrence risks specific to *NFIX* and similar genes remain sparse. This gap in the literature underscores the need to expand the list of genes associated with elevated recurrence risk, even when the precise pathomechanisms remain unexplored. Including such genes in recurrence risk assessments could significantly improve genetic counseling and risk prediction for affected families.

These observations highlight the importance of considering the specific gene involved during genetic counseling. When a *de novo* variant is identified in a gene with documented evidence of gonadal mosaicism in fathers - such as *NFIX* in the context of Malan syndrome - genetic counselors should inform couples about the potential recurrence risk. To increase the accuracy of risk assessment, analyzing the most informative and accessible tissue, such as the father’s semen, using highly sensitive detection methods for low VAFs is essential. Our dataset, which includes two fathers exhibiting semen-only mosaicism, demonstrates that analyzing the paternal germline is crucial for detecting mosaicism that may not be evident in less targeted samples, such as blood or buccal swabs. This approach enhances the accuracy of recurrence risk assessments and genetic counseling, as it helps identify cases where low VAFs may be missed otherwise.

Phasing of *de novo* variants in our study confirms a 3:1 – 4:1 ratio of paternal origin, consistent with previous findings^4,46^. By analyzing semen samples for low-level mosaicism in cases with paternal *de novo* variants and using a detection threshold of 0.2%, we can stratify recurrence risk for affected couples. If paternal gonadal mosaicism is excluded by analysis of semen sample, the recurrence probability is negligible. Conversely, if mosaicism is confirmed, the recurrence risk can be estimated based on the VAF of the detected mosaicism, enabling refined genetic counseling. As demonstrated by the family of IND_027 in our cohort, which includes two affected children and the lowest confirmed VAF in the paternal semen sample, it is essential to have comprehensive discussions with each family regarding the implications for optional prenatal diagnostics. The presence of low-level paternal mosaicism can complicate recurrence risk assessments, requiring careful counseling to help families make informed decisions.

Maternally arising *de novo* variants are more challenging, as the germline tissue is not accessible. Testing multiple tissues will allow the detection of maternal mixed mosaicism with commonly a moderate VAF (two cases in our cohort with 7-23%). Taken into account that paternal *de novo* events are by far more prevalent, we do not expect that there will be mosaic variants in the ovaries that cannot be detected in other tissues. This is however speculative and needs to be proven.

Considering the literature and our results, we now suggest the following management when parents of a child with a *de novo* variant ask about the recurrence risk (Figure 5).

**Figure 5.**
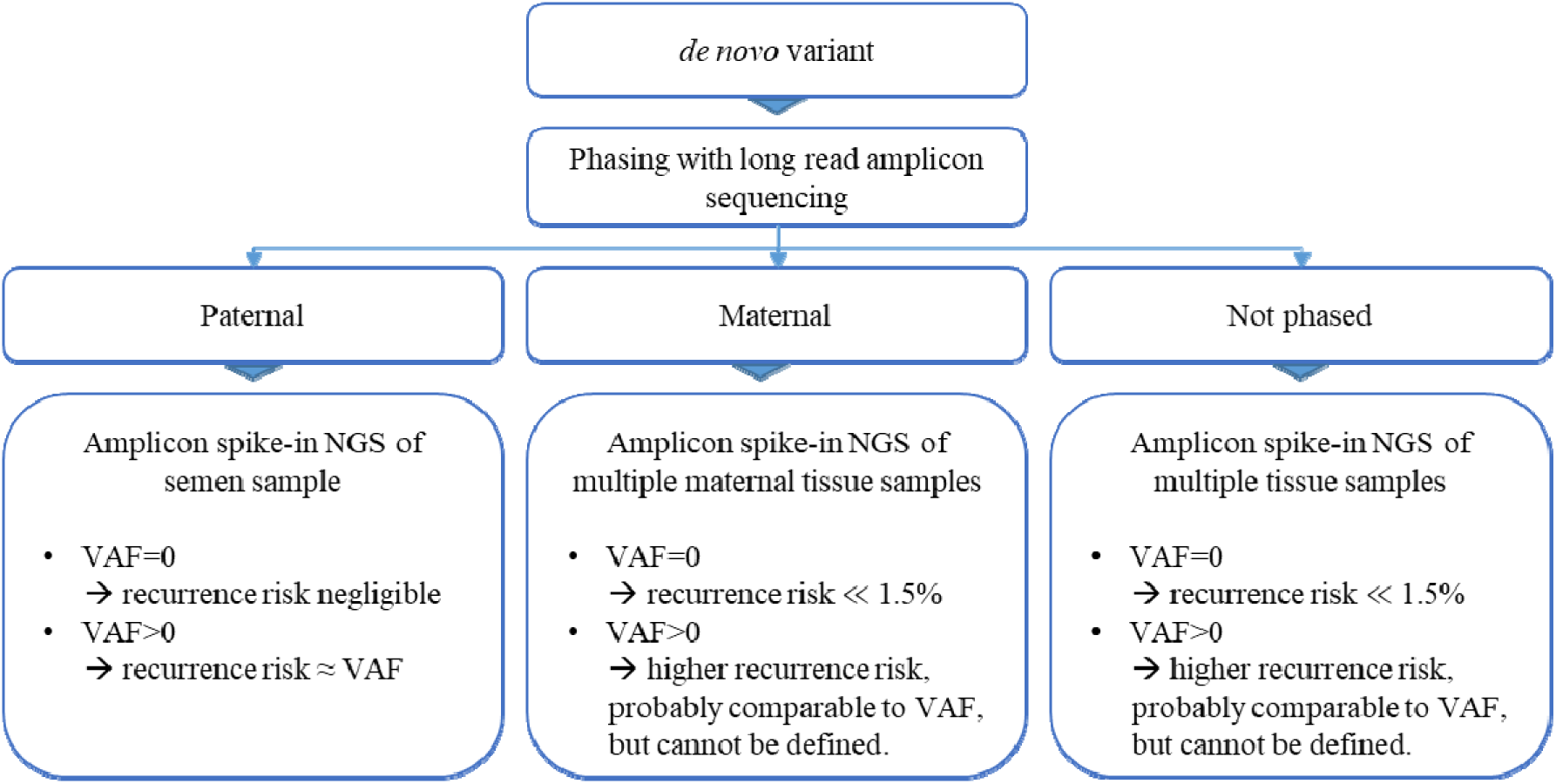
Flowchart of recommended analyses for personalized counselling on recurrence risk for a specific de novo variant.

The general risk that one of the parents carries a mosaic in any tissue is 7.5% (overall weighted mean of our meta-analysis).

– To personalize this risk, first, phase the variant (Table S1 in Zenodo^22^, sheet “primer_phasing”).
– In about ¾ to 4/5 of the cases, the variant is paternal. Then, analyze the semen sample. If the variant can be excluded as mosaic in the semen, recurrence risk can be considered as negligible. If there is a mosaic, recurrence risk can roughly be given as equivalent to the VAF. However further testing, e.g. prenatal diagnostics, has to be discussed depending on the needs of the family.
– If the variant is on the maternal allele, analyze multiple tissues of her, e.g. blood, buccal swab, and hair follicle. If the variant can be excluded as mosaic in all samples, there is a remaining probability that only an ovary carries the variant. We consider this as very low. In men, we observe 1.5% of mosaic only in semen and further 1.5% in semen and in other tissues. Naïve transfer to women, who show similar number of mixed mosaicism (1.5%), means that approx. further 1.5% have a gonadal mosaic in the ovary and not in other tissue. This is fully not plausible when considering the parental origin of the *de novo* variants (only ¼ to 1/5 are maternal). Therefore, we may consider this as the maximum estimation, while the truth is probably much lower.

This management is possible case per case as our experiments show that a simple amplicon of the variant and spiking this in an NGS run leads to reliable and comparable results.

## Conclusions

Our aggregated results highlight that parental mosaicism in cases of *de novo* variants in offspring is more common than previously assumed, with VAFs varying considerably between tissues. This variability underscores the limitations of assigning a generalized recurrence risk of 1–5% in clinical practice. Instead, risk estimation can and should be refined through advanced methods, such as variant phasing and targeted spike-in PCR combined with high-depth NGS analysis. While broader studies are needed to fully understand the prevalence and impact of parental mosaicism, it is increasingly clear that recurrence risk must be tailored not only to individual cases but also to specific genes. Incorporating these approaches will enhance the accuracy of genetic counseling and support more informed reproductive decision-making for affected families.

## Data Availability

All data produced in the present study are available upon reasonable request to the authors

https://zenodo.org/records/15769707

## Abbreviations

MAP: Maximum A-Posteriori estimate
VAF: Variant Allele Fraction
NGS: Next-Generation Sequencing
UMI: Unique Molecular Identifiers

## Declarations

## Author contributions

Sonja Neuser: Conceptualization, Data curation, Methology, Visualization, Writing – Original Draft. Natalie Ahmad: Resources, Project administration. Data Curation. Denny Popp: Formal analysis, Writing – Original Draft. Julia Hentschel: Resources. Johannes Lemke: Resources, Knut Krohn: Resources, Anna-Lisa Schween: Resources, Maike Karnstedt: Writing – Original Draft, Stephan Drukewitz: Data curation, Methodology, Writing – Original Draft, John Wiedenhöft: Formal analysis, Methodology, Writing – Original Draft. Rami Abou Jamra: Conceptualization, Supervision, Writing – Original Draft. All Authors: Writing - Review & Editing.

## Data and code availability

All data generated or analyzed during this study can be found in the online version of this article at the publisher’s website.

## Supplemental Tables

Table S1: Information of clinical data and variants in the cohort. Methodical information on panel design and primers for phasing and validation.

Table S2: Literature for meta-analysis.

Table S3: Variant allele fractions of all analyzed variants before UMI collapse, after UMI collapse and maximum a-posteriori of variant allele fraction from Bayesian model.

Neuser, S. (2025). Table S1-3. (Zenodo). https://doi.org/10.5281/ZENODO.15769707 https://doi.org/10.5281/ZENODO.15769707.

